# Clinical evaluation of a RT-LAMP SARS-CoV-2 test for the Point-Of-Care, rapid, low-cost, integrating sample solid phase extraction and on which reagents are lyophilized

**DOI:** 10.1101/2021.10.03.21264480

**Authors:** E. Coz, P. Garneret, E. Martin, D. F. do Nascimento, A. Vilquin, D. Hoinard, M. Feher, Q. Grassin, J. Vanhomwegen, J.C Manuguerra, S. Mukherjee, J.-C. Olivo-Marin, E. Brient-Litzler, M. Merzoug, E. Collin, P. Tabeling, B. Rossi

**Affiliations:** MMN, CBI, ESPCI, 6 rue Jean Calvin, Paris 5, France; Gulliver CNRS UMR 7083, PSL Research University, ESPCI Paris, 10 rue Vauquelin, 75005 Paris, France; CIBU, Institut Pasteur, 25-28 rue du Dr Roux, Paris 15; Bioimage Analysis Unit, Institut Pasteur, 25-28 rue du Dr Roux, Paris 15^3^; CNRS UMR 3691, 25-28 rue du Dr Roux, Paris 15; Innovation Office, Institut Pasteur, 25-28 rue du Dr Roux, Paris 15; CMC Ambroise Paré, 25 Boulevard Victor Hugo, 92200 Neuilly sur Seine; Hôpital Ballanger, Blvd Robert Ballanger, 93600 Aulnay-Sous-Bois

## Abstract

**Objectives:** Determine the sensitivity and specificity of a Point-Of-Care test (‘COVIDISC’) for SARS-COV2. The novelty of the test is to integrate, on the same (low-cost) compact plastic/paper device, solid phase RNA extraction and RT-LAMP amplification, all reagents being freeze-dried on it.

**Method:** Retrospective study with a cohort of 99 patients characterized by real-time RT-PCR. The 37 positive naso-pharyngeal samples cover a broad range of viral loads (from 5 gc /µL to 2 10^6^ gc/ µL of sample).

**Results:** The COVIDISC found 36 positives (out of 37 by IP4 RT-PCR protocols) and 63 negatives (out of 62 by RT-PCR).

**Conclusion:** The sensitivity of the COVIDISC, found in this 99-patient retrospective study, is 97% and the specificity 100%.

## Introduction

Over the last eighteen months, a number of spread models of COVID have been developed (See for instance [1][2][3][4]). They represent valuable inputs for elaborating efficient testing strategies. One question of interest, addressed in these models, and much relevant technologically, is the influence of testing frequency and test sensitivity on the spread rate mitigation of the disease. RT-PCR is the most sensitive test [5], but, being time-consuming and costly, it cannot be performed at high frequencies and thus may miss a significant number of contaminated cases. On the other hand, antigen tests (such as BinaxNow), are much less sensitive [6], but being low cost and simpler to deploy, can be performed more frequently. Models proposed different trade-offs. From these modelling studies, one may suggest, although no quantitative analysis has been done yet, that the development of new molecular tests, as sensitive as RT-PCR, and whose costs and simplicity of use are comparable to antigen tests, could substantially alleviate the situation. In this spirit, a number of nucleic acid amplification tests (NAAT) have recently been proposed [7], following up an effort initiated one decade ago [8]. Most of these NAATs are based on isothermal amplification, in particular Loop-Mediated Isothermal Amplification (LAMP), for which, now, a large documentation is available [9]. RT-LAMP reaction takes place at constant temperature (65°C) and thus does not need thermocycling machines [10][11]. However, to reach performances comparable to the gold standard (RT-PCR), nucleic acid extraction is required. Today, extraction is traditionally made in spin column-based RNA extraction. The process, involving centrifugation and a number of pipetting steps, is difficult to accommodate with the low-cost Point of Care (POC) constraints. To circumvent the difficulty, several RT-LAMP tests, targeting the POC market, have reduced sample preparation to heating or chemical treatment [12][13][14][15]. These simplifications are indeed interesting from a POC viewpoint, but in all cases, and unsurprisingly, they led to a drop of the sensitivity [16]. By standing significantly below the gold standard, and slightly above the antigen test, the question of the competitive positioning of these tests in the diagnostic landscape, is raised.

Here, we develop a new molecular Point-Of-Care RT-LAMP test, that has the potential to improve significantly or change the situation. The main novelties are the integration of a solid phase extraction on the device and the lyophilization, on it, of all components needed to perform RT-amplification. The idea is that solid phase extraction (with fluids driven by capillarity forces, instead of centrifugal forces) allows to reach high sensitivities, while freeze-drying facilitates transportation and storage. In a recent work [17], we tested the concept on a laboratory all-paper system. Since then, we engineered a compact device, called COVIDISC, in which we modified the design and the way how fluids are manipulated, keeping the biological process the same and the cost at a low level. Here we investigate the clinical performances of a series of prototypes of this device. The goal of the present work is thus to assess the clinical performances of this new test, i.e. its sensitivity and specificity, based on a cohort of patients spanning a broad range of viral loads.

## Materials and Methods

### COVIDISC description

The COVIDISC consists of two plastic disks, 8.5 cm in diameter, able to rotate around a common axis (see Fig 1). The device performs SARS-CoV-2 RNA detection (Orf1a/b gene)[18] and human 18S RNA detection, thus integrating a positive control [19]. Silica membranes, cut in form of small disks, are placed in lodgings located in the upper plastic disk. These membranes are dedicated to perform nucleic acid extraction. They are called ‘capture membranes’. A large absorbing pad is placed on the lower plastic disk. It is dedicated to pull, by capillarity, the fluids through the capture membranes, during the extraction step. A removable funnel guides the fluids and inhibits contamination. In another lodging, a capsule contains two additional membranes, called detection or reaction membranes (or disks). In them, the RT-LAMP reagents are freeze-dried. The time-life of the lyophylisate is several months (data not shown). One lyophylisate includes the SARS-CoV-2 RT-LAMP mix, deposited on the circular membrane, and the other contains the human 18S RNA RT-LAMP mix, deposited on the square-shaped membrane. Both membranes are placed in a removable capsule (see Suppl Figure 1A). The description of the RT-LAMP reaction mix composition and the freeze-drying process can be found in [17]

**Figure 1:**
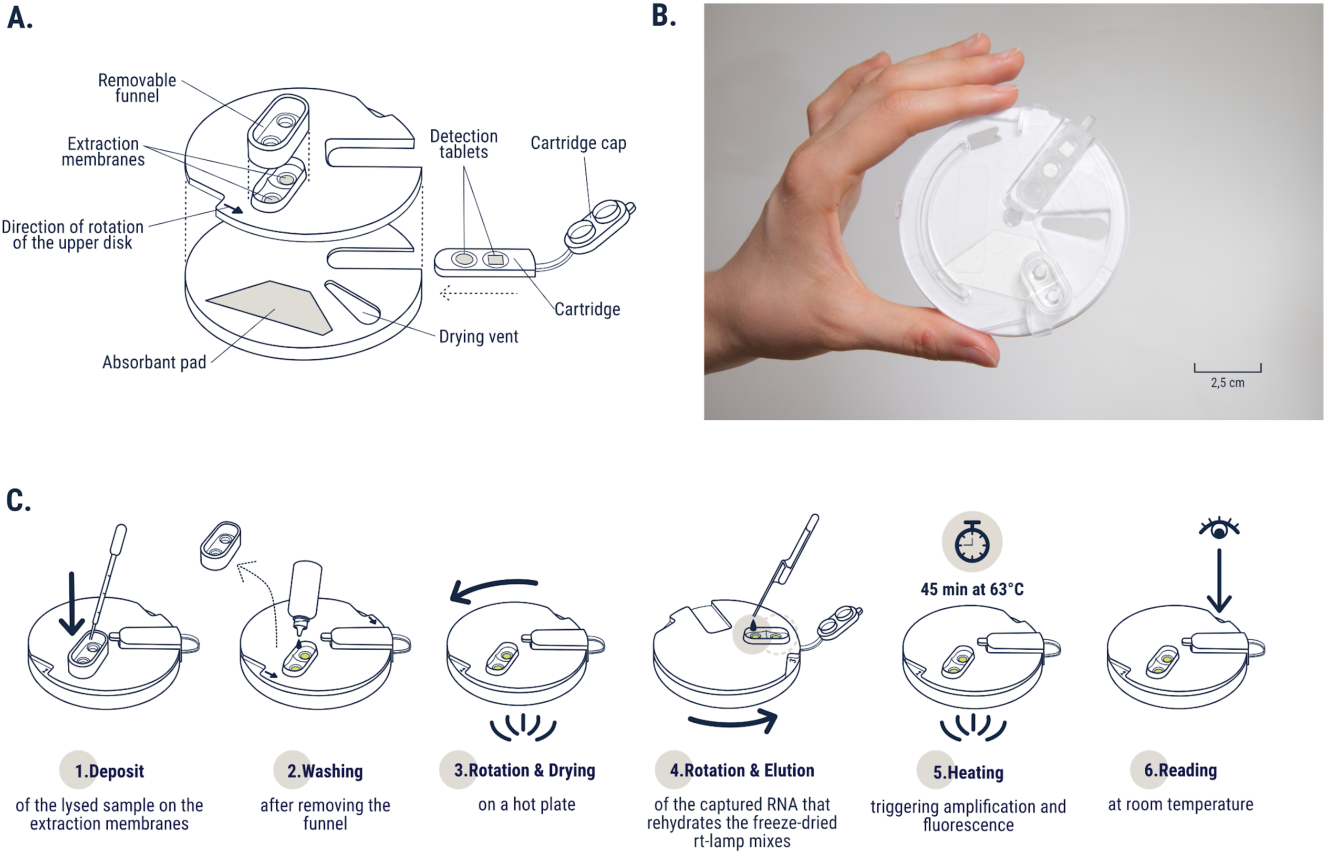
(**A)** Exploded view of the COVIDISC **(B)**. COVIDISC photograph. (**C)**. COVIDISC workflow : 1 – Sample lysis at 65°C before sample injection. 2 – Washing of the extraction membranes. 3 – Upper disk rotation and drying at 65°C. 4 – Upper disk rotation and RNA elution. 5 – Upper disk counterrotation and closing of the cartridge. 6 – 45 min heating and readout at room temperature.

### COVIDISC Fabrication

The COVIDISC is manufactured by thermoplastic injection molding (Protolabs, France). The lower part of the COVIDISC and the removable funnel are made of white polypropylene. The cartridge and the upper part of the COVIDISC are made of transparent polypropylene. The insertion of the capture membranes and reaction membranes in done manually

### COVIDISC Protocol

The workflow is shown in Figure 1: after being mixed, at 65°C, with a chaotropic lysis buffer, the sample is injected in the extraction unit, i.e. the two extraction membrane, placed in contact with the pad, through a funnel that guides the fluids and inhibits wall contamination. After the sample injection is achieved, the funnel is removed, and the membranes are rinsed with 400µL of a 70% ethanol solution, further dried at 65°C for 15 min. Then the disc is rotated, bringing the two extraction or capture membranes (one for the sample, the other for the positive control), in contact with the two reaction membranes. The nucleic acids captured in the extraction membranes are eluted into the reaction membranes, using an aqueous solution. The eluates, driven by capillarity, imbibe the reaction membranes, and thus hydrate the freeze-dried RT-LAMP reagents. After heating at 65°C for 45 minutes, the capsule is removed, placed on a visualization device, composed of a blue LED screen and an orange filter (Blue Light Transilluminatorsc) and imaged with a low-cost USB camera (Dino-Lite). To avoid issues raised by the colorimetric phenol-red LAMP products (acidic conditions of samples leading to false positives) [20], we coupled our RT-LAMP reaction to a specific fluorescent-probe-based method developed by Ball et al. [21], called QUASR. A simple fluorescent reader and a smartphone camera are then sufficient for reading the result of the test. With the sample and buffer manipulations included, a total time of, approximately, one hour is needed to perform the test.

### Image analysis

In order to discriminate the positives from the negatives, we developed two algorithms. In algorithm 1, we define, for each image of the reaction membrane, a quantity equal to the average to the 1% highest intensity levels. By comparing this quantity to the background intensity, we declare the test positive or negative. We checked that the results were not critically dependent of the fraction of percentiles we choose, within a range between 0.1% and 5%. This algorithm, based in the intensity levels, attempts to mimic the direct naked-eyes readout, which is possible with our system, thanks to the high fluorescence intensity of the positive cases, provided by the QUASR technique. Data obtained with Algorithm 1 is shown in Fig 2, the detail being given in Supplementary Materials, ‘Image analysis with Algorithm 1’). In Algorithm 2, we define a Covidness scores by analyzing the intensity level distributions emitted by each reaction membrane. This method, which emphasizes more on the gradients of intensity than Algorithm 1, and designed to be more robust, will be described in a further paper.

**Figure 2:**
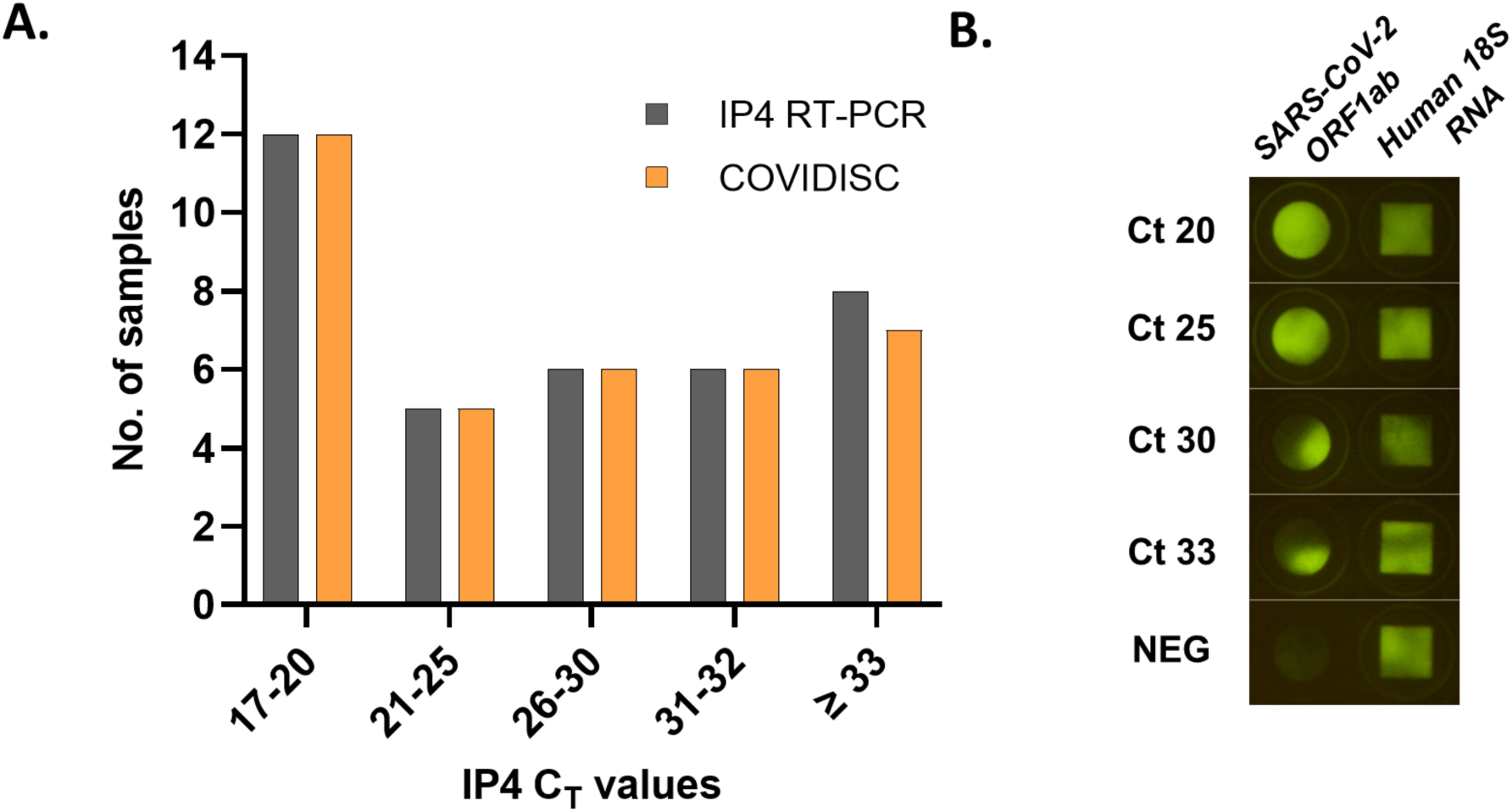
<u>A</u>: <u>Comparison between COVIDISC and IP4 RT-qPCR [22]</u>: Histogram of the tests declared positive by PCR (dark gray bars) and COVIDISC (orange bars), as a function of the cycle threshold (Ct) values provided by IP4-qPCR [22].<u>B: Images of COVIDISC reaction zones after amplification</u> (i.e. approximately one hour after the sample is introduced in the device). The squares are the positive controls (18S human RNA) and the disks are the samples. CT values, obtained by IP4 RT-qPCR, are indicated. At large viral loads (CT = 20 and 25) the fluorescence signal is spatially homogeneous. At low viral load (CT=30 and 33) the fluorescent signal is localized in spots.

### Clinical samples

Clinical samples consist of 100 naso-pharyngeal swabs resuspended in Universal transport medium (UTM, Copan 330C, VPM Improviral™ and VTM Sigma-Virocult®). Those samples are obtained from hospitalized patients or from patients consulting emergencies in a period extending from 30^th^ March 2020 and 14^th^ August 2020 (period with a positivity rate between 20 and 50 %) at Robert Ballanger Hospital and stored at −80°C at the day of collection. 100 samples were randomly chosen from the entire collection. One sample was excluded because the resuspension volume was below 200µL. To compare with the gold standard, RT-PCR of the 99 samples were performed at CNR (Center National of Reference), using a protocol described in [22]. The RT-qPCR analysis showed that the 99 clinical samples included 37 positive samples, with RT-PCR cycle threshold values (Ct) ranging from 18 to 33 for the targeted RNA-dependent RNA polymerase *RdRP* gene region (designated as “IP4”). The viral load associated with the CT value of 33 was estimated to be close to 5 genome copies/µL of sample, based on a quantified standard (Vircell amplirun MBTC030).

### Clinical trial

The 99 COVIDISC were prepared a few days before the clinical trial. In practice, the trial, performed at Robert-Ballanger hospital, took one day. The 99 samples were defrosted at 4°C. Two 200µL aliquots were prepared for each sample. 8 COVIDISC were simultaneously run.

### Ethical statement

The study was approved by National Institute of Health and Medical Research (INSERM) Ethics Evaluation Committee, the Institutional Review Board (IRB00003888).

### Characterization of the analytical Sensitivity and Specificity of the COVIDISC on nasopharyngeal and saliva matrices

The analytical limit of detection (LOD) of the COVIDISC was determined by using a quantified control (Vircell amplirun MBTC030), in which inactivated viral particles of known concentrations were spiked in a nasopharyngeal matrix. The LOD was estimated to 3.2 genome copies per µl (gc/µl) (see Supplementary Figure 1A). The specificity against a series of viruses, on all-paper systems, using the same workflow as the COVIDISC, was 100% [17]. Similar characterizations, carried out in nasopharyngeal (Supplementary Figure 1B) and saliva matrices (Supplementary Figure 1C) led to sensitivities equal to 3.5 ± 1.5 genome copies per µl (gc/µl) of sample.

## Results

Fig 2A shows the essential result of our work, i.e. the comparison between COVIDISC and RT-PCR. On the figure, the CT values of the 37 positive samples, obtained by IP4 RT-PCR, span from 17 to 33, i.e. from 2.10^6^ to 5 gc/µl. Important to note, the set of samples shown in Fig 2A includes a substantial number of low viral loads. To establish Fig 2A, we used Algorithm 1, whose results are consistent with the naked eye. COVIDISC detects 36 positive samples out of 37, leading to a clinical sensitivity of 97.3 %. The false negative is close to our analytical LOD (Ct 33, around 5 gc/µL); this probably explains why we did not detect it. The remaining 62 samples, declared negative by real-time RT-PCR, were also found negative by the COVIDISC, leading to a specificity of 100%.

To provide more detail on the test, Fig 2B shows the fluorescence emitted by the reactive disk, after amplification, for positive samples of various viral loads (CT values of 20, 25, 30 and 33) and, for the sake of comparison, one negative sample. The RNA 18S positive control (square-shaped) are unambiguously positive. In the disks (which contain the samples), and in the positive cases, the fluorescence signals are sufficiently strong be visualized with the naked eye. The samples with the largest viral loads (associated to RT-PCR CT values of 20 and 25) produce an emission of fluorescence homogeneously spread on the reaction disk area. For samples with the lowest viral loads (associated to RT-PCR CT values of 30, 33), the fluorescence is still high, but it is localized inside spots occupying a fraction of the disk area. This fraction decreases with the viral loads. We hypothesize that the phenomenon is due to a nucleation process: when the viral load is small, a small number of RNA strands are present in the reaction disk (we estimated this number between 10 and 30 at CT=33), and, consequently, the spreading of the reaction throughout the entire area gets subjected to statistical uncertainty. In all cases, whether spotty or homogeneous, the detection of the positives can be clearly done, either by eye, or by algorithm 1. Table 1 summarizes the results of our study, in which the statistical errors, due to the limited size of the cohort, are calculated.

**Table 1.** Comparison between COVIDISC and (IP4) RT-PCR [22].

| **IP4 Ct Category** | **COVIDISC (%, 95% CI)** | **Total** |
| --- | --- | --- |
| **Total positive** | 36 (97.3, 85.8-99.9) | 37 |
| **Low (>30)** | 14 (93.3, 68.1-99.8) | 15 |
| **Medium (20-30)** | 16 (100, 79.4-100) | 16 |
| **High (<20)** | 6 (100, 54.1-100) | 6 |
| **Negative** | 62 (100, 94.2-100) | 62 |

## Conclusion

To summarize, the retrospective clinical study reported here shows that the COVIDISC clinical sensitivity is 97% (85.8-99.9) and its specificity is 100% (94.2-100). In brief, what we show here is that COVIDISC performs as well as RT-PCR platforms. We may add that the device integrates extraction and amplification, uses reagents freeze-dried on the device, and is low cost (the production cost, reagents included, is estimated to 5 €). Important to note, it could also be used with saliva (see Supplementary Figure 2C). These characteristics suggest that COVIDISC could be deployed on a much larger scale than the gold standard (RT-PCR), and, in the meantime, offer a sensitivity orders of magnitude better than antigen tests (in terms of viral load). Although quantifying the impact of such a test, in a manner shown in [2], has not been made yet, one may suggest that COVIDISC conveys, at least potentially, an opportunity to bend the curves of the kinetics of contamination in a more effective manner than the existing technologies.

## Data Availability

The data we refer to is included in the paper and in the references.

## Author information

PT, E.Coz, PG, E.Collin, BR and MM designed the study. E.Coz, PG, EM and DGdN prepared the COVIDISCS. E.Coz, PG, DH, MF, QG and E.Collin, performed the study. E.Coz, PG, SM, J-CO,EBL and A.V analyzed the data. PT, E.Coz and PG wrote the manuscript. EBL, JV, JCM edited the manuscript.

## Transparency declaration

No conflict of interest.

## Acknowledgment

The authors thank La Ville de Paris, ESPCI, CNRS, PSL, its Department of Tech Transfer “PSL Valorisation”, IPGG platform (ANR-10-EQPX-34), the Institut Pasteur, the institut Carnot Pasteur Maladies Infectieuses (ANR-11-CARN-017-01), the Region Ile-de-France DIM ELICIT program, the Labex IBEID (ANR-10-LABX-62-IBEID), the France BioImaging infrastructure (ANR-10-INBS-04), ANR Flash, Fonds de l’ESPCI, Bettencourt Foundation, JL Missika, MC Lemardeley and T.Coulhon for their support. They thank the members of MMX group at IPGG, K. Bala, V. Croquette, P.Vitaux, C. Brechot, L.Dehove for discussions and helps.

## Supplementary material

### Analytical sensitivity with nasopharyngeal and saliva matrixes

**Supplementary Figure1:**
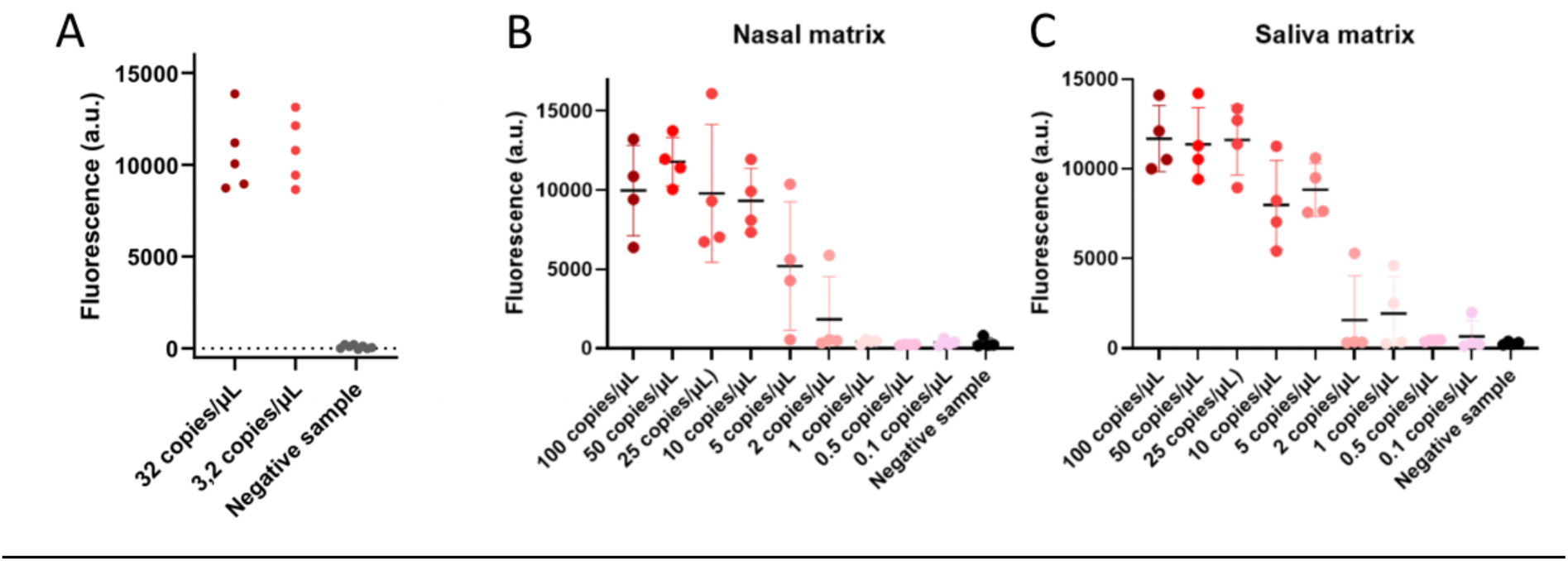
Analytical characterizations of the COVIDISC. A - Extraction and amplification, on COVIDISC, of SARS-CoV 2 calibrated samples (Vircell amplirun MBTC030). The total fluorescence intensity within the circle-shaped reaction area is represented. Limit of detection is < 3.2 gc/µL sample for naso-pharyngeal samples are obtained. B – COVIDICS intensity measurements of inactivated virus samples spiked in nasopharyngeal matrix, at various concentrations. C – Same as B, but in a saliva matrices.

### Image analysis with Algorithm 1

**Supplementary Figure 2:**
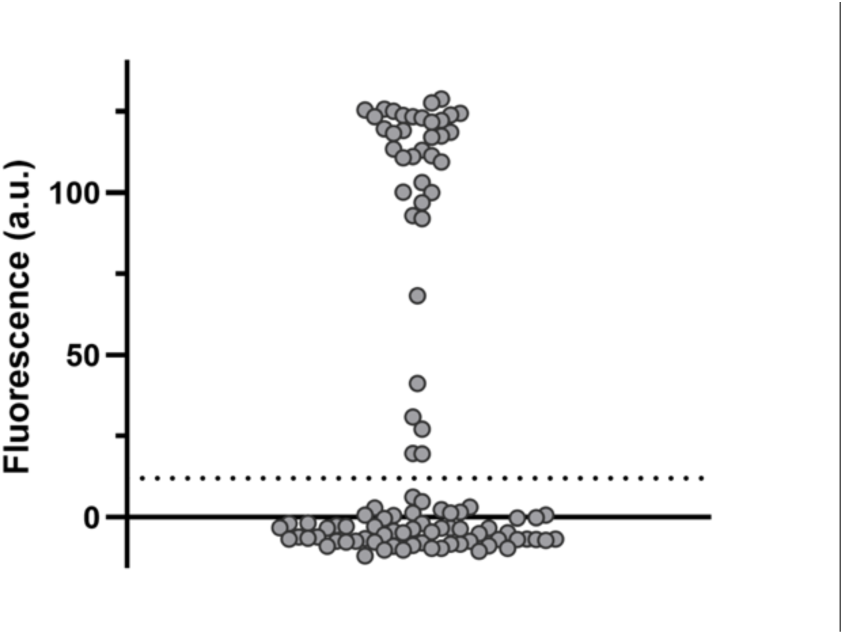
Algorithm 1: Distribution of fluorescence intensities (averaged intensities of top 1 percentile), of the 99 samples, after amplification. The abscissa is chosen to separate the data. The ordinate is the grey level determined by the camera. The dashed line, situated at a grey level equal to 12, is determined in a way that it substantially lies above the background. This line clearly separates the negatives (below 12) from the positives (above 12). The ratio, in terms of grey levels, between the darkest positive and the brightest negative is equal to 4. Thereby, positive and the negative samples can be distinguished without ambiguity.

## References

[1] Mina, M.J, Roy Parker M.D, Larremore D.B., « Rethinking Covid-19Test Sensitivity - A Strategy for Containment - », N. Engl. J. Med., p. 383:e10, 2020.

[2] Forde, J.E., Ciupe, SM, « Quantification of the Tradeoff between Test Sensitivity and Test Frequency in a COVID-19 Epidemic - A Multi-Scale Modeling Approach», Viruses, p. 457, 2021.

[3] Choi W., Shim E., « Optimal strategies for social distancing and testing to control COVID-19», J.Theor.Biology, vol. 512, p. 110568, 2021.

[4] Vassallo L., Perez I.A., Alvarez-Zuzek L.G., Amaya J., Torres, M.F., Valdez L.D., La Rocca C.E., Braunstein L.A., et Vassallo L., Perez I.A.,Alvarez-Zuzek L.G., J. Amaya G.J., Torres M.F., Valdez L.D., La C.E., « An epidemic model for COVID-19 transmission in Argentina: Exploration of the alternating quarantine and massive testing strategies», Math. Biosciences, p. https://doi.org/10.1016/j.mbs.2021.108664, 2021.

[5] World Health Organization, « Diagnostic testing for SARS-CoV-2: interim guidance, 11 September 2020 (No. WHO/2019-nCoV/laboratory/2020.6). »

[6] Perchetti GA, Huang ML, Mi MG, Jerome KR, Geninger AL, « Analytical sensitivity of the Abbott BinaxNOW COVID-19 Ag card», J Clin Microbiol, vol. 59, p. e02880–10, 2021.

[7] Chaouch, M., « Loop mediated isothermal amplification (LAMP): An effective molecular point of care technique for the rapid diagnosis of coronavirus SARS CoV 2», Rev Med Virol, p. e2215, 2021.

[8] Magro L, Escadafal C, Garneret P, Jacquelin B, Kwasiborski A, Manuguerra JC, Monti F, Sakuntabhai A, Vanhomwegen J, Lafaye P, Tabeling P., « Paper microfluidics for nucleic acid amplification testing (NAAT) of infectious diseases», Lab. Chip, vol. 17, p. 2347–71, 2017.

[9] Becherer, L, Nadine Borst N, Bakheit M., Frischmann S., Zengerle R., von Stetten F., « Loop-mediated isothermal amplification (LAMP) – review and classification of methods for sequence specific detection», Anal Methods, vol. 12, p. 717, 2020.

[10] Carter, L.J., et al, « Assay Techniques and Test Development for COVID-19 Diagnosis», ACS Cent Sci, vol. 6, p. 591-605, 2020.

[11] Esbin, M.N., et al., « Overcoming the bottleneck to widespread testing: a rapid review of nucleic acid testing approaches for COVID-19 detection », vol. 26, p. 771–783, 2020.

[12] L’Helgouach, N., Champigneux, P., Santos-Schneider, F., Molina, L., Espeut, J., Alali, M., …, Molina, « EasyCOV: LAMP based rapid detection of SARS-CoV-2 in saliva», MedRXiv, 2020.

[13] Fowler, V. L., Armson, B., Gonzales, J. L., Wise, E. L., Howson, E. L., Vincent-Mistiaen, Z.,… & Kidd, S. P., « A reverse-transcription loop-mediated isothermal amplification (RT-LAMP) assay for the rapid detection of SARS-CoV-2 within nasopharyngeal and oropharyngeal swabs at Hampshire Hospitals NHS Foundation Trust», MedRXiv, 2020.

[14] Schellenberg, J. J., Ormond, M., Keynan, Y., « Extraction-free RT-LAMP to detect SARS-CoV-2 is less sensitive but highly specific compared to standard RT-PCR in 101 samples.», Journ Clin Vir, p. 104764, 2021.

[15] Gangulia, A, Mostafa A, Bergera J. Aydinc MY, Sunb F., Stewart de Ramireze A, Valera Cunninghama, BT, King WP, Bashir R., PNAS, vol. 117, p. 22727–22735, 2020.

[16] Taki, K., Yokota, I., Fukumoto, T., Iwasaki, S., Fujisawa, S., Takahashi, M., Negishi, S., Hayasaka, K., Sato, K., Oguri, S., Nishida, M., Sugita, J., Konno, S., Saito, T., Teshima, T., « SARS-CoV-2 detection by fluorescence loop-mediated isothermal amplification with and without RNA extraction.», J. Infect. Chemother. Off. J. Jpn. Soc. Chemother., vol. 27, p. 410–412, 2021.

[17] Garneret P., Coz E., Martin E., Manuguerra JC., Brient-Litzler E., Enouf V., González Obando D.P., Olivo-Marin JC., Monti F., van der Werf S., Vanhomwegen J., Tabeling P., « Performing point-of-care molecular testing for SARS-CoV-2 with RNA extraction and isothermal amplification.», Plos One, vol. 16, p. e0243712. 7., 2021.

[18] Lamb, L. E., Bartolone, S. N., Ward, E., Chancellor, M. B., « Rapid detection of novel coronavirus/Severe Acute Respiratory Syndrome Coronavirus 2 (SARS-CoV-2) by reverse transcription-loop-mediated isothermal amplification.», Plos One, vol. 8, p. 1–9, 2018.

[19] Lamb, L. E., Bartolone, S. N., Ward, E., Chancellor, M. B., « Rapid detection of Zika virus in urine samples and infected mosquitos by reverse transcription-loop-mediated isothermal amplification», Sci. Rep., vol. 8, p. 1–9.

[20] Yang, Q., Meyerson, N. R., Clark, S. K., Paige, C. L., Fattor, W. T., Gilchrist, A. R., Sawyer, S. L., « Saliva TwoStep for rapid detection of asymptomatic SARS-CoV-2 carriers.», MedRXiv, 2021.

[21] Ball, C. S., Light, Y. K., Koh, C. Y., Wheeler, S. S., Coffey, L. L., Meagher, R. J, « Quenching of unincorporated amplification signal reporters in reverse-transcription loop-mediated isothermal amplification enabling bright, single-step, closed-tube, and multiplexed detection of RNA viruses», Anal Chem, vol. 88, p. 3562–3568.

[22] World Health Organization, « Protocol: real-time RT-PCR assays for the detection of SARS-CoV-2, Institut Pasteur, Paris. World Health Organization, Geneva. Available via https://www.who.int/docs/default-source/coronaviruse/real-time-rt-pcr-assays-for-the-detection-of-sars-cov-2-institut-pasteur-paris.pdf », 2020.

